# Are there significant correlations between climate factors and the spread of COVID-19 for less densely populated and less polluted regions?

**DOI:** 10.1101/2021.02.11.21251129

**Authors:** Marcelo de Paula Corrêa, Ana Letícia Campos Yamamoto, Luiz Felipe Silva, Ivana Riêra Pereira Bastos, Tális Pereira Matias, Raquel Gonçalves Pereira, Patrícia Martins do Reis, Flávia Fernanda Azevedo Fagundes, Alysson Fernando Ribeiro, Joaquim Augusto Cortez Moraes, Filipe Pereira da Silva

## Abstract

This study analyzes the correlation between the spread of COVID-19 and meteorological variables (air temperature, relative humidity, wind speed, and precipitation) in urban-rural cities located in southeastern Brazil. Spearman’s correlation coefficients were used for the statistical analysis. Results show that air temperature and wind speed were positively correlated with COVID-19 cases, while air relative humidity showed negative correlation. As seen in several recent studies, climate factors and the spread of COVID-19 seem to be related. Our study corroborates this hypothesis for less densely populated and less polluted regions. We hope that our findings help worldwide scientific efforts towards understanding this disease and how it spreads in different regions.

**Highlights:**

- Climate and COVID-19’s spread were also correlated in less-densely populated regions.
- Both maximum and minimum temperatures are strongly correlated with cases of covid-19.
- One hypothesis for the strong association could be the high minimum temperatures in the subtropical region.
- Wind speed is also positively correlated with COVID-19, while air humidity is negatively related.
- Mitigation policies against the spread of COVID-19 should be based on local climate profiles.

## 1. Introduction

On January 30, 2020, the World Health Organization (WHO) declared a Public Health Emergency of International Concern (PHEIC) due to the rapid spread of the new coronavirus disease (COVID-19) around the world (WHO, 2020; Ye et al., 2020; Lai et al., 2020). This disease is caused by severe acute respiratory syndrome coronavirus 2 (SARS-CoV-2) and it was first reported in Wuhan, China in December 2019 (Zhu et al., 2020a).

COVID-19 is mainly spread by human-to-human contact via respiratory droplets or small airborne particles produced by an infected individual (Li et al., 2020). In other words, transmission can occur via secretions (saliva) or respiratory droplets (PAHO/WHO, 2020). Although most people who contract the disease develop mild symptoms, it can cause severe respiratory problems and other collateral health problems in others, and this effect is still being studied (England et al., 2020; He et al., 2020). In addition to demographic and social factors, the transmission of SARS-CoV-2 can be influenced by meteorological conditions such as temperature, humidity, wind speed, radiation, air pollution, and precipitation (Bukhari et al., 2020; Rosario et al., 2020; Zhang et al., 2020). Thus, airflow is an important factor in the spread of COVID-19 (via aerosols), similar to other diseases (Shakil et al., 2020), and meteorological variables may influence this process.

Recent studies have shown that weather is an important factor in determining the rate of COVID-19 transmission (Şahin, 2020; Tosepu et al., 2020; Bashir et al., 2020). In fact, meteorological variables may be more relevant in predicting mortality rates compared to other variables, such as population, age, and urbanization (Malki et al., 2020). Initially, it was thought that virus transmission would be greater in cold climates. However, COVID-19 transmission is observed everywhere in the world, including areas with hot and humid weather. Brazil, with its tropical climate, is one of the worst affected countries in the world with the third highest number of COVID-19 infection cases. Almost 195,000 deaths were caused by Covid-19 in 2020, according to a panel from the Brazilian Ministry of Health (MS, 2020) and data from December 31.

Since studies on weather data and COVID-19 cases can offer important information on the behavior of the virus, and offer potential improvements for controlling the spread of the disease (Auler et al., 2020), we investigated possible correlations among data collected from southeastern Brazil. Unlike most studies published on the topic, this focuses on studying urban-rural small cities.

## 2. Methods

### 2.1. Study area

Brazil is a tropical country with high temperatures. However, climatic conditions may vary drastically due to the country’s size (8.5 million of km^2^). The northern regions in Brazil are mostly tropical with Savanna (As), Monsoon (Am) and Rainforest (Af) climates, according Köppen’s classification. On the other hand, the southern regions have milder climates, with cold winters and hot summers. In the southeastern regions of Brazil, there is marked seasonal rainfall, with humid summers and dry winters (Cf, Cs and Cw, according to the Köppen classification) (Ramos et al., 2018; Martins et al., 2018).

The study area concentrated on one of 12 mesoregions in the south/southwest region of the state of Minas Gerais (MG), which covers 146 cities and towns (IBGE, 2020a) (Figure 1). The annual average temperature is 20°C influenced by topographic variations with altitudes ranging from 76 to 2892 m (INMET, 2020; Oliveira et al., 2018; Reboita et al., 2015). This mesoregion has the second highest economic index in the state, with high living standards and good urban infrastructure (Cirino and González, 2011). We selected a sample of six cities due to the abundance of meteorological data, the number of hospitalizations, and the climatic conditions (Table 1).

**Table 1:** Demographic and geographic information on the study region.

| City | Acronym | Latitude and Longitude | Area (km<sup>2</sup>) | Altitude (m) | Population * |
| --- | --- | --- | --- | --- | --- |
| Itajubá | ITJ | -22.4°; -45.4° | 294.8 | 856 | 97,334 |
| Machado | MAC | -21.7°; -45.9° | 586.0 | 880 | 42,413 |
| Maria da Fé | MdF | -22.3°; -45.4° | 202.9 | 1258 | 14,056 |
| Passa Quatro | PAQ | -22.4°; -45.0° | 277.2 | 938 | 16,393 |
| São Sebastião do Paraíso | SSP | -20.1°; -47.0° | 814.9 | 980 | 71,445 |
| Varginha | VAR | -21.5°; -45.4° | 395.4 | 980 | 136,602 |

Source: IBGE (2020b)
\*Estimated population (2020)

**Fig. 1.**
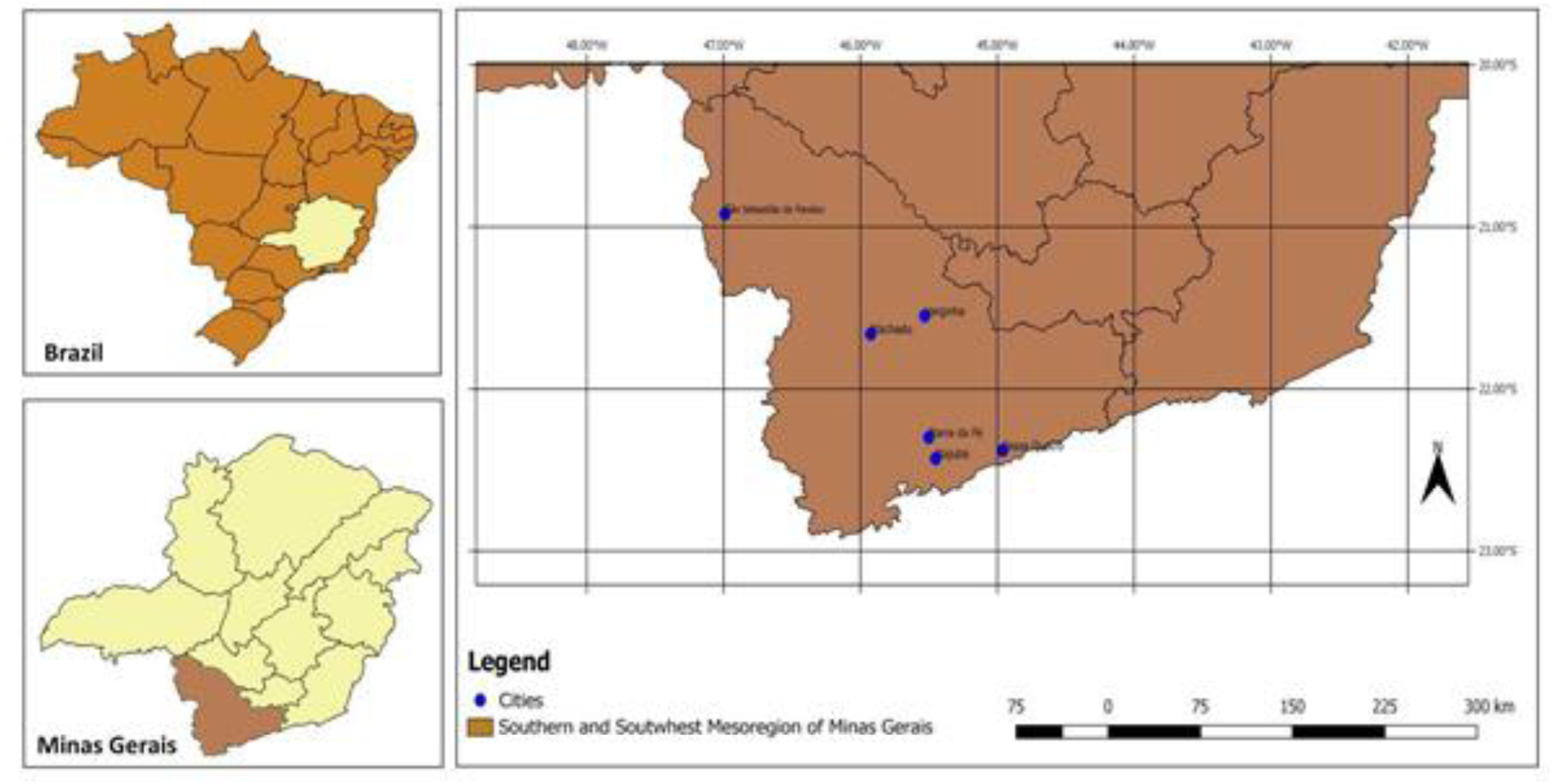
Geographic location of the study region.

### 2.2 Data collection

Meteorological data were taken from automatic weather stations managed by the National Institute of Meteorology (INMET, 2020) and the Federal University of Itajubá (UNIFEI, 2020). The data was taken for daily minimum temperature (Tmin – °C), maximum temperature (Tmax – °C), average temperature (T – °C), relative humidity (RH – %), wind speed (WS - m/s), and daily accumulated rainfall (DAR – mm) for the period between April and December 2020. Data on the number of COVID-19 cases were obtained from the database of Secretary of Health of the Government of Minas Gerais state (SHGMG, 2020).

### 2.3. Data analysis

SigmaPlot 12.5 software (Systat Software Inc., USA) was used to calculate Spearman’s correlation coefficient for the six study locations. We investigated the correlation of meteorological data and daily cases of COVID-19, from the first case until August 31, 2020. Our sample covered different time periods for the different cities: ITJ (261 days), MAC (269 days), MdF (229 days), PAQ (232 days), SSP (258 days), and VAR (269 days).

## 3. Results and Discussi

### 3.1 Meteorological data

Figure 2 shows the boxplots of Tmin, Tmax, and RH. Mean Tmax ranged from 24.1 ± 3.2°C in MdF to 28.5 ± 3.6°C in SSP, and mean Tmin from 9.9 ± 4.0°C in MdF to 12.9 ± 3.4°C in PAQ. MdF, is a city located at a relatively high altitude, and therefore registered lower temperatures, sometimes as low as −1.7°C during the winter. During the winter, atmospheric circulation contributes to reduced moisture in the region (Martins et al., 2018). Less than 10% of RH were lower than 40%. Most of measurements were observed in winter. Despite dry winters, the 1^st^ quartile of RH was greater than 56% for all cities. However, RH can decrease to 20% or 30% in the afternoons of drier days. Mean WS, not shown in Figure 2, was around 1 to 2 m/s, and maximum mean daily values were close to 10 m/s. Wind gusts were not accounted for in the dataset.

**Fig. 2.**
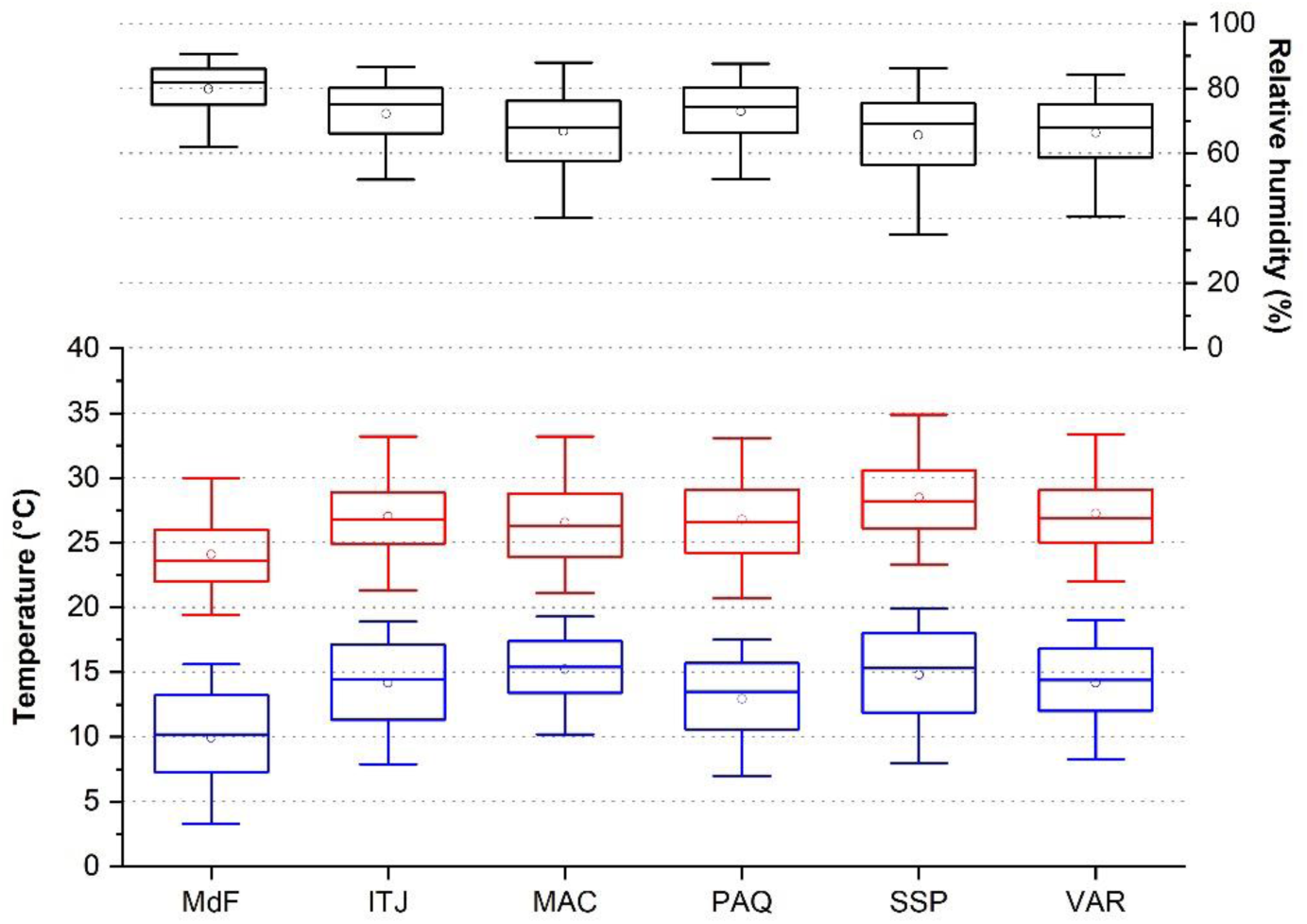
Boxplot of some meteorological variables for the six studied locations between April and December, 2020. Blue: Minimum temperature (°C), Red: Maximum temperature (°C), and Black: Relative humidity (%). Vertical rectangles depict the range of the first and third quartiles; horizontal solid lines are the median values; whiskers limit percentiles P5 and P95; and circles represent the averages.

Seasonal behavior of rainfall in Southeast Region of Brazil is seen in Figure 3. Winter is the dry season, while summer is the rainy season. Even though all of the cities are located in the same region, the accumulated DAR values varied significantly. MdF had the highest accumulated DAR, followed by ITJ and MAC. More than 90% of the annual rainfall is observed in the summer, when storms are frequent with DAR about 70 to 80 mm day-1. On the other hand, clear-sky days are typical in June and July, as there is little to no rain in the period. According to the National Institute of Meteorology, 2020 had normal precipitation accumulation (near climatological mean).

**Fig. 3.**
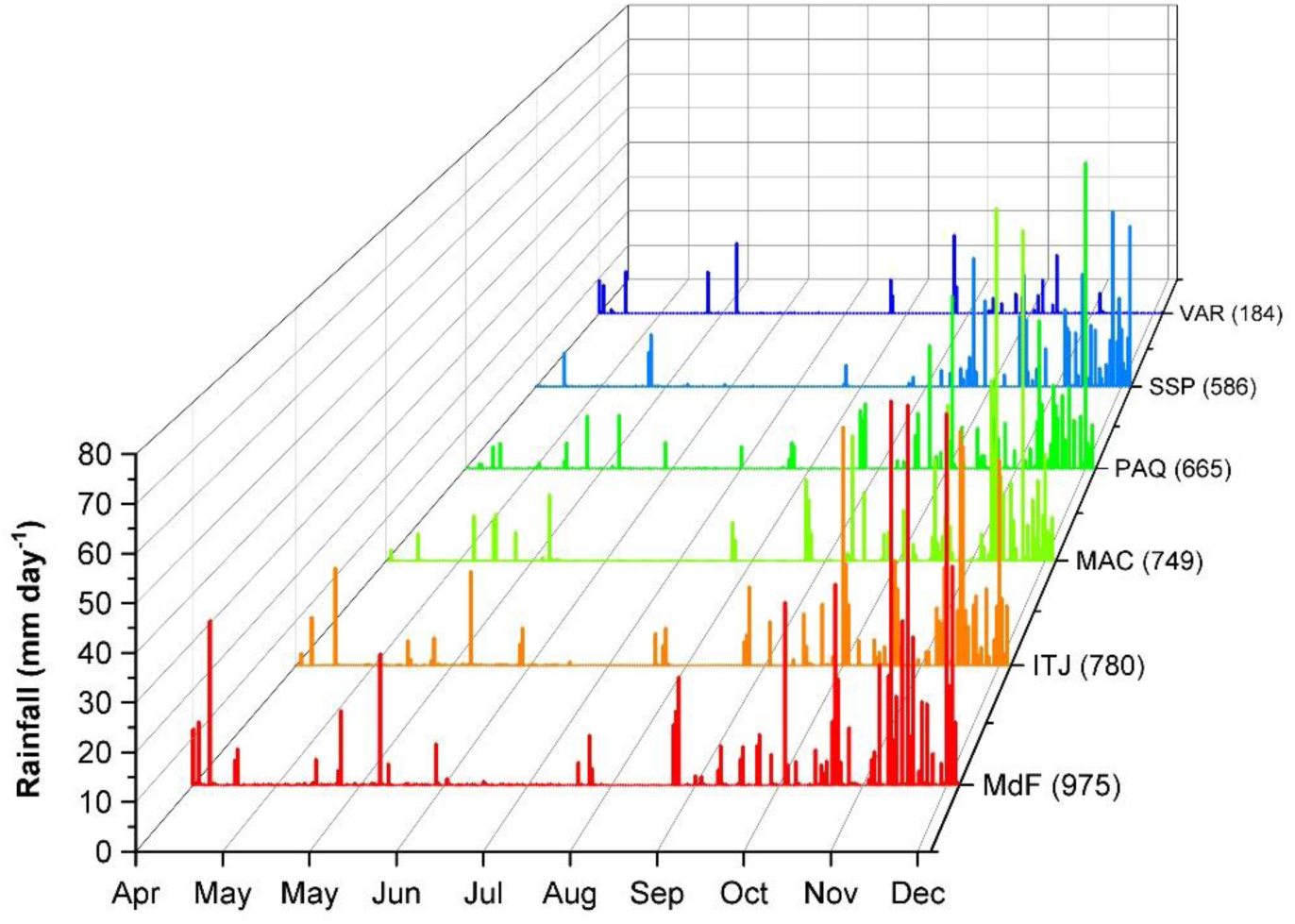
Rainfall (mm day^-1^) between April and December, 2020. The values in parentheses show the accumulated rainfall (mm) in the period

### 3.2 COVID-19 spread

Figure 4 shows the reported cases of COVID-19 per city. In the six cities evaluated, 6,694 cases were confirmed in 2020, a cumulative incidence of 1781 cases per 100,000 inhabitants. The cumulative incidence of cases in the region is lower than that observed in the Minas Gerais state (2565/100,000) and half of what was observed in Brazil (3653/100,000). ITJ, VAR and SSP registered more than 87% of all cases. However, the latter shows the largest incidence with 2335 cases per group of 100,000 inhabitants.

**Fig. 4.**
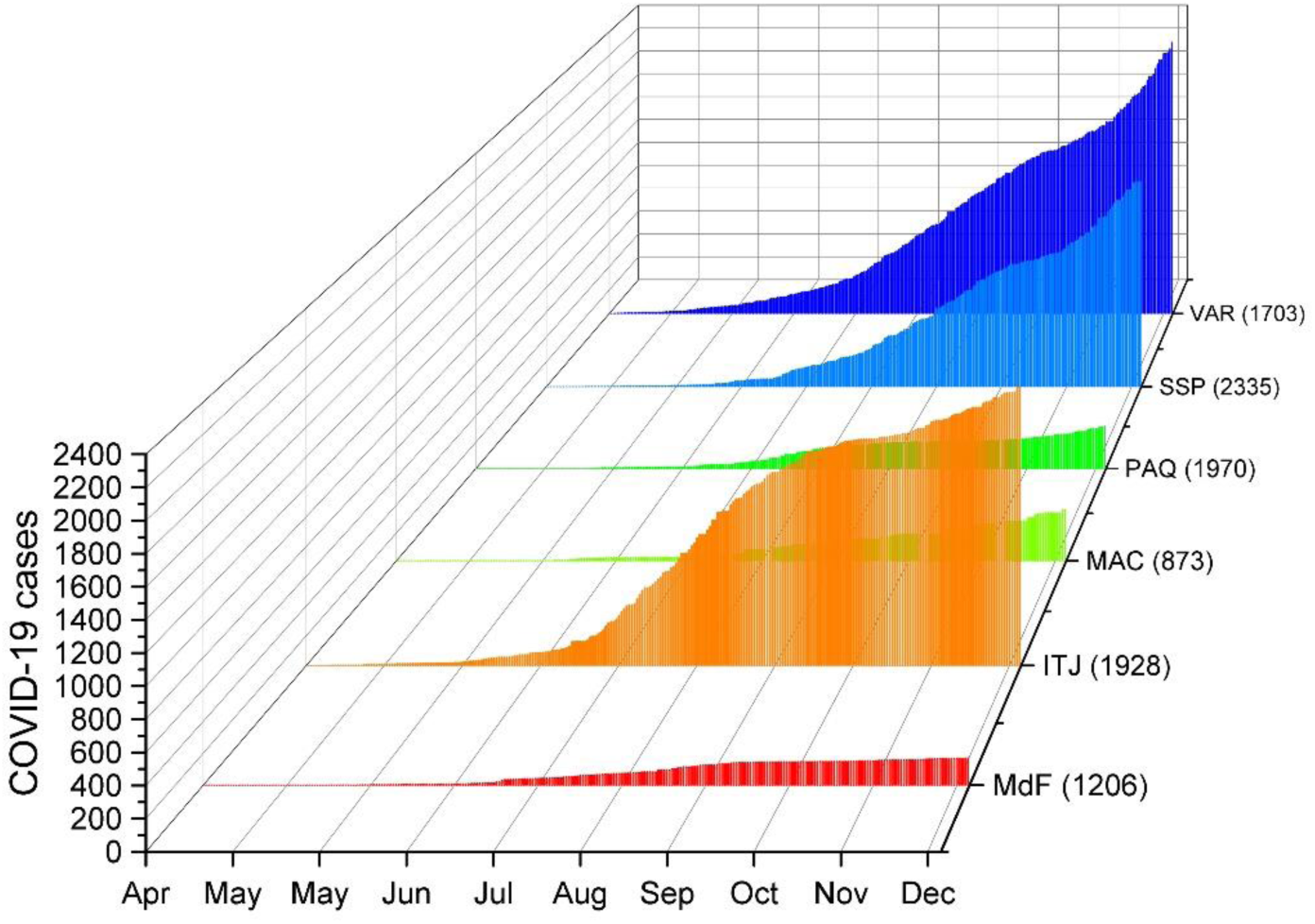
COVID-19 confirmed cases in 2020 in cities studied. Values in parenthesis show the cumulative incidence data of cases per 100,000 inhabitants.

### 3.3 Correlation between COVID-19 and meteorological variables

Spearman’s correlation coefficients between meteorological variables and the daily cases of COVID-19 are shown in Table 2. T followed by RH showed stronger correlation with COVID-19 cases. On the one hand, Tmax, Tmin and Tmean were positively correlated with COVID-19 incident in almost all cities. On the other hand, RH showed significant inverse correlations in most of them. WS and COVID-19 cases showed positive correlation in three of the six cities. Finally, DAR did not show any correlation with disease’s cases incidence.

**Table 2:**
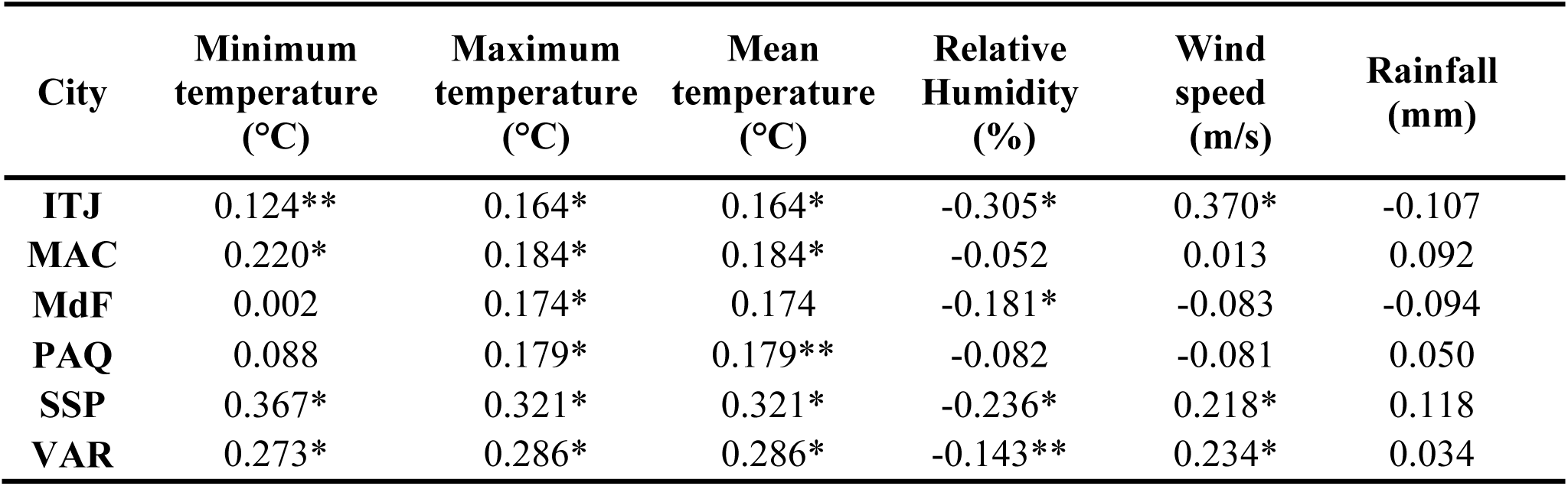
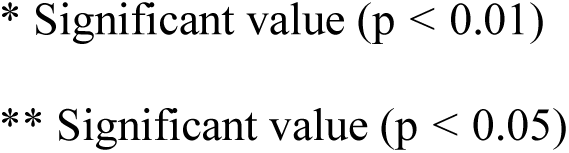
Spearman correlation coefficients between COVID-19 and weather variables

| City | Minimum temperature (°C) | Maximum temperature (°C) | Mean temperature (°C) | Relative Humidity (%) | Wind speed (m/s) | Rainfall (mm) |
| --- | --- | --- | --- | --- | --- | --- |
| ITJ | 0.124** | 0.164* | 0.164* | -0.305* | 0.370* | -0.107 |
| MAC | 0.220* | 0.184* | 0.184* | -0.052 | 0.013 | 0.092 |
| MdF | 0.002 | 0.174* | 0.174 | -0.181* | -0.083 | -0.094 |
| PAQ | 0.088 | 0.179* | 0.179** | -0.082 | -0.081 | 0.050 |
| SSP | 0.367* | 0.321* | 0.321* | -0.236* | 0.218* | 0.118 |
| VAR | 0.273* | 0.286* | 0.286* | -0.143** | 0.234* | 0.034 |

\* Significant value ( $p < 0.01$ )
\*\* Significant value ( $p < 0.05$ )

## 4. Discussions and Conclusion

This study investigated the association between meteorological variables and registered cases of COVID-19 in low populated cities located in the southeastern region of Brazil. The main goal was to evaluate if the correlations observed in other recent studies are also significant for less densely populated and less polluted regions. In the first year of pandemic, the incidence of cases in the region of study was almost the half of than those observed in Brazil. One preliminary hypothesis for the lower incidence could be the better air quality in small cities.

RH is a significant factor in the survival and transmission of viruses in indoor environments (Casanova et al., 2010; Chan et al., 2011; Paynter, 2014; Ahlawat et al., 2020) and low RH contributes to increases in viral respiratory infections (Souza and Sant’Anna Neto, 2008; Gardinassi et al., 2012). Significant correlations between RH and virus transmission rates were verified in New York USA (Bashir et al., 2020) and 30 Chinese provinces (Qi et al., 2020). Both studies showed that RH and T have significant adverse impacts on new cases of COVID-19. Zhu et al. (2020b) also found a highly significant correlation between daily incubated cases and absolute humidity (calculated from the air temperature and relative humidity) at eight South American locations. Since previous studies have shown associations between air pollution and negative impacts on human health (Wolkoff; Kjærgaard, 2007; Leitte et al., 2009; Wolkoff, 2018), we might expect weaker associations between RH and COVID-19 at urban-rural locations. However, our study shows that COVID-19 contamination was strongly linked to RH even in less polluted environments.

Tmax and Tmin showed positive correlations for different locations, respectively. A recent study performed in Brazil showed that high temperatures (27.5 °C) and RH (~80%) positively influenced COVID-19 transmission rates (Auler et al., 2020). In Jakarta, high mean temperatures (26.1 to 28.6 °C) were correlated with COVID-19 (Tosepu et al., 2020). Sharma et al. (2020) and Menebo (2020) also found similar results on positive correlations between Tmax and COVID-19 transmission. In our study, the highest significant positive correlations for Tmax (r = 0.367; p < 0.01) and Tmin (r = 0.321; p < 0.01) were found in SSP. Significant associations were not observed in coldest cities, MdF and PAQ. In summary, our results indicate that increased T levels and decreased RH levels seem to be associated to increased COVID-19 cases. This correlation pattern is similar to the previous studies performed in larger cities (Auler et al. (2020); Bashir et al. (2020)).

WS showed significant correlation with COVID-19 incidence in 3 of 6 cities. Unlike previous study recently performed in Rio de Janeiro (Rosario et al., 2020), our results indicate positive correlation between WS and disease’s cases. In general, WS disperses pollutants in the atmosphere and stronger winds should result in less contamination (Afiq et al., 2012; Zhang et al., 2019). However, WS vary significantly over geographic locations and time. Furthermore, wind speed can vary greatly from coastal regions to the interior, and so the comparisons with the results presented in the Rio de Janeiro study need more detailed evaluation. Indeed, further studies are needed to assess how the virus spreads in function of outdoor wind weather conditions.

DAR and COVID-19 were not correlated in our study. A recent study performed in Oslo, Norway Menebo (2020) showed significant correlation between lack of precipitation and increased COVID-19 cases in. Perhaps the lack of statistical significance in our study is associated with the related to the discrepancies in the rainfall regime between the driest season (winter) and wettest season (summer) in the region.

This study seeks to contribute to helping the scientific community understand, and therefore, mitigate the spread of COVID-19 and contain the pandemic. Recent studies have already shown how weather conditions impact the spread of the virus, and increased knowledge on the matter can contribute to employing preventive actions. Thus, our contribution was focused on analyzing the relationship between weather conditions and COVID-19 in smaller cities. Public policy and governmental action should take local climate characteristics into account (Shakil et al. 2020). Furthermore, we suggest that future studies focus on the long-term weather impacts on the spread of COVID-19. These studies should also consider associations based on information relating to comorbidity, social distancing rates, air pollution, vulnerability, and socioeconomic factors (Shen et al., 2021; Srivastava, 2021).

## Data Availability

All data is available. Please contact the main author.

## Acknowledgements

A. L. C. Yamamoto would like to thank the Coordination for the Improvement of Higher Education Personnel (Coordenação de Aperfeiçoamento de Pessoal de Nível Superior – CAPES) for their financial support. Finance Code 001. We also thank Federal University of Itajuba for the financial support.

